# Melodic Intonation Therapy for aphasia: A multi-level meta-analysis of randomised controlled trials and individual participant data

**DOI:** 10.1101/2021.08.28.21262764

**Authors:** Tudor Popescu, Benjamin Stahl, Brenton M. Wiernik, Felix Haiduk, Michaela Zemanek, Hannah Helm, Theresa Matzinger, Roland Beisteiner, W. Tecumseh Fitch

## Abstract

Melodic Intonation Therapy (MIT) is a prominent rehabilitation programme for individuals with post-stroke aphasia. The present meta-analysis investigated the efficacy of MIT while considering outcome measure quality, experimental design, influence of spontaneous recovery, MIT protocol, and level of generalisation.

An extensive literature search identified 606 studies in major databases and trials registers; of those, 22 studies, overall 129 participants, met all eligibility criteria. Multi-level mixed- and random-effects models served to separately meta-analyse RCT and non-RCT data.

RCT evidence on validated measures revealed a small-to-moderate standardised effect in non-communicative language expression for MIT, with substantial uncertainty. Unvalidated measures attenuated MIT’s effect size compared to validated tests. MIT’s effect size was 5.7 times larger for non-RCT data compared to RCT data. Effect size in non-RCT data decreased with number of months post-stroke, suggesting confound through spontaneous recovery. Variation from the original MIT protocol did not systematically alter benefit from treatment. Progress on validated tests arose mainly from gains in repetition tasks rather than other domains of verbal expression such as everyday communication ability.

The current results confirm the promising role of MIT in improving trained/untrained performance with unvalidated measures, alongside validated repetition tasks; whilst highlighting possible limitations in promoting everyday communication ability.

## 1. Introduction

Stroke survivors often experience a profound loss of communication skills, among them a syndrome known as aphasia. This syndrome may manifest as severe difficulty in verbal expression, referred to as ‘non-fluent aphasia.’ In addition, stroke survivors frequently suffer from impaired speech-motor planning. Known as ‘apraxia of speech,’ this syndrome often occurs in combination with aphasia. Although about a third of individuals with neurological communication disorders do not recover completely,^1^ rehabilitation programmes can improve language performance even in the chronic stage of symptoms.^2^

Melodic Intonation Therapy (MIT) is a prominent rehabilitation programme originally developed for individuals with non-fluent aphasia.^3^ Drawing on the observation that individuals with neurological communication disorders are often able to sing entire pieces of text fluently,^4–6^ MIT uses melody, rhythm, vocal expression in unison and alone, left-hand tapping, formulaic and non-formulaic verbal utterances, as well as other therapeutic elements in a hierarchically structured protocol.^7^ Hypotheses on MIT’s neural mechanisms have been discussed.^8^

To date, randomised controlled trial (RCT) data have confirmed the efficacy of MIT on validated outcome measures in the *late subacute* or *consolidation* stage of aphasia (i.e., up to 12 months after stroke),^9^ but not in the *chronic* stage of symptoms (i.e., more than 6–12 months after stroke).^10^ From a methodological perspective, influences of spontaneous recovery are generally lower in the *chronic* stage of aphasia, as suggested by RCT data^11^ and meta-analyses.^12^ This points out the need carefully to consider stage of symptoms post-stroke in research on MIT. Importantly, speech-language therapy seeks to promote performance on untrained items. Consistent with this goal, the present work distinguishes progress on *trained* items—that is, learning resulting from using the same set of utterances both during treatment and subsequent assessment—from the more desirable goal of attaining generalisation to *untrained* items, ideally in the context of everyday communication to ensure ecological validity.^13^

So far, there have been several systematic reviews^14,15^ and meta-analyses on MIT.^16–18^ Existing meta-analyses reflect a relatively limited amount of RCT data,^16^ dichotomise post-treatment improvement in a way that prevents specific effect size estimates^17^ or do not operationalise quality of outcome measures (psychometrically validated vs. unvalidated tests), experimental design (presence vs. absence of randomisation and control group), influence of spontaneous recovery (quantified as number of months post-stroke), MIT protocol applied (original vs. modified), and level of generalisation (performance on trained vs. untrained items).^18^ Given the substantial burden of disease associated with aphasia, the current meta-analysis attempts to provide a deeper understanding regarding the clinical potential and possible limitations of MIT. To achieve this goal, our analyses synthesise available studies on MIT to address five research questions focusing on whether the effect size of the rehabilitation programme is systematically altered by:

1. **Psychometric quality**: use of validated vs. unvalidated tests in outcome measures;
2. **Experimental design**: RCT vs. non-RCT studies;
3. **Confound by spontaneous recovery**, decreasing with number of months post-onset of stroke (MPO);
4. **Deviation in protocol**: original vs. slightly modified MIT variants;
5. **Degree of generalisability**: performance on trained vs. untrained items.

## 2. Methods

### 2.1. Eligibility criteria

We defined the following basic **inclusion criteria** for primary studies to be considered for the present meta-analysis:

1. empirical study with or without a control group that administered MIT to adult individuals with aphasia (aged at least 18 years);
2. language-related outcomes in pre-post assessment.

We chose to include case reports with individual participant data (IPD) to increase the pool of evidence. To determine the influence of experimental design on treatment outcome, we analysed RCT and non-RCT studies separately and comparatively.

After removal of duplicate items (see Supplementary Materials, §1), the following **exclusion criteria** were applied to remaining studies (in chronological order):

1. publication in non-peer-reviewed or predatory journal;
2. unvalidated outcome measure (i.e., no published or otherwise accessible work confirming the psychometric properties of a particular test battery). Exceptionally, if a study included both trained and untrained items for an unvalidated measure, we included this work to determine the degree of generalisation by comparing performance on trained and untrained items;
3. other essential data not reported and / or not retrievable, even after contacting the authors (e.g., no sample size or standard error, insufficient information to compute an effect size);
4. substantial variation from original MIT protocol.^3^ We accepted *minor* changes to the MIT protocol (and examined the effect of the categorical variable: original *vs*. modified MIT), as long as the modified protocol had all of the following features:
  i. melody-based vocal expression both in unison and alone;
  ii. some form of rhythmic pacing (e.g., left-hand tapping);
  iii. use of verbal utterances known from everyday communicative interaction.
  Aside from the original version of MIT, seven modified MIT protocols were reported across studies initially considered before applying our protocol-related exclusion criteria. Applying these exclusion criteria resulted in four MIT protocols finally included (citations indicate the first description of the protocol itself, where available, or studies employing it):
  i. Modified Melodic Intonation Therapy (MMIT)^19^;
  ii. Singing, Intonation, Prosody, “Atmung” (German for “breathing”), Rhythm, and Improvisation (SIPARI)20;
  iii. Speech-Music Therapy for Aphasia (SMTA)^21^;
  iv. “singing therapy”^22,23^.
  Excluded protocols, with reasons for exclusion, were:
  i. Metrical Pacing Technique (no melodic intonation alongside rhythmic pacing);
  ii. aphasia choirs / choir therapy (unison singing only; use of regular song lyrics rather than verbal utterances known from everyday communicative interaction);
  iii. music therapy combined with SLT (no melody-based vocal expression by patients taken individually).

Taken together, the included studies comprised 129 treated participants (59 in RCTs; 70 in IPDs) and 62 control participants (all in RCTs). The full list of included and excluded studies can be found in eTable 1 and eTable 2 (Supplementary Materials).

### 2.2. Search strategy

To obtain high search sensitivity, we used both free-text and subject headings in databases for our search, not restricting by language or publication form.^24^ Figure 1 shows the PRISMA statement chart (Preferred Reporting Items for Systematic Reviews and Meta-Analyses^25^), which summarises the study counts at all stages of the search. The full counts are given in the Supplementary Materials, §1, which also documents the full literature search procedure, including search terms, databases used, and attempts made to reach the “grey literature”.

**Figure 1:**
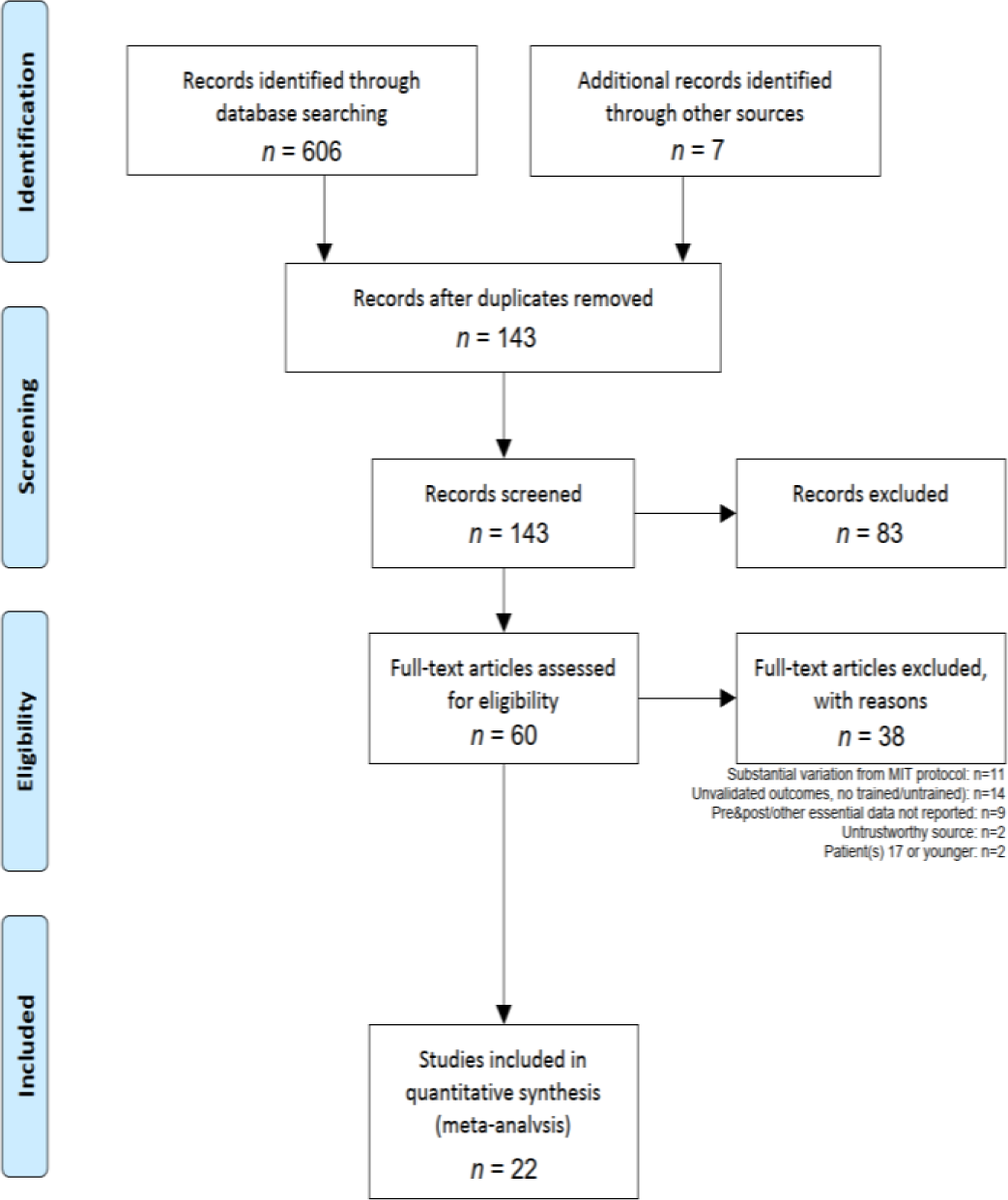
Flow diagram from the PRISMA statement (http://prisma-statement.org/prismastatement/Checklist.aspx). The lists of included and excluded studies are in eTables 1 and 2 of the Supplementary Materials, respectively.

Furthermore, we followed the guidelines and standards in the Methodological Expectations of Cochrane Intervention Reviews (MECIR) handbook, and those in the PRISMA checklist (see Supplementary Materials).

### 2.3. Study coding and double - coding

All studies were coded by the first author (TP). Two of the authors (FH, TM) re-coded all studies, verifying the cross-coder consistency. Agreement among the three coders occurred in a majority of cases, and any discrepancies found between coding sheets were solved by consensus. The ICCs (intraclass correlations) were >0.9 in the remaining cases, which amounted to errors arisen from numerically estimating data reported in plot format only.

### 2.4. Tests and outcome measures in primary studies

All test batteries reported in the studies considered, and their validation status, are reported in eTable 3 of the Supplementary Materials. eTable 4 *ibid*. shows in detail which of the subtests of these test batteries are measuring which linguistic Ability; the associated Target Syndrome (aphasia or apraxia of speech); and the hierarchical categorisation scheme that determined in each case the dependent variable meta-analysed (Domain). An abridged, tree-form version of this categorisation scheme (not showing test batteries and their subtests) is shown in Figure 2.

**Figure 2:**
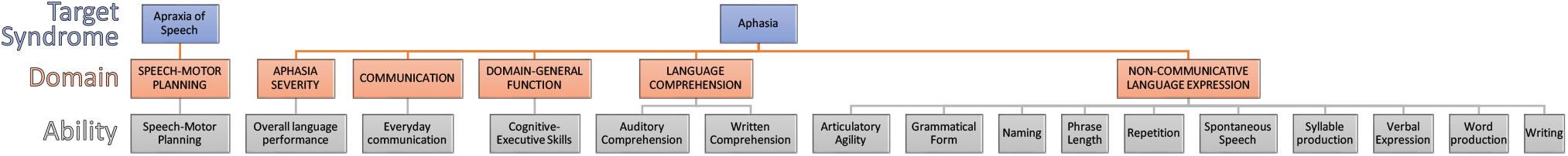
Hierarchical categorisation scheme. Shows how Abilities nest into Domains, within each Target Syndrome level considered. All meta-analyses were done at the Domain level of abstraction.

### 2.5. Meta - analysis methods

#### 2.5.1. Computed outcome metric

To maximise comparability of effects across studies, we used change scores from pre-test to post-test as the outcome metric, expressed in *z*-scores. For group-level studies (the RCTs in the current analyses), we standardised *z*-scores using pooled pre-test standard deviation across control and treatment groups. For IPD studies (the case reports in the current analyses), we computed *z*-scores in one of three ways. For studies that reported results as *z*-scores (e.g., based on test norms), we used the *z*-scores directly. For studies that reported results as percentile scores (e.g., based on test norms), we converted these to *z*-scores using the quantiles of the standard Normal distribution. For other studies, we estimated *z*-scores using the following procedure: We first converted^a^ normalised raw scores to reflect the proportion of the maximum possible score, POMP.^26^ Next, we estimated a three-level random-intercept model for the pre-test POMP scores, with individual test scores nested within patients nested within studies (see Figure 3). From these models, we used the population intercept as the estimated POMP score *mean*, and the patient-level random effects standard deviation as the estimated POMP score *SD* (τ). We then used this *mean* and *SD* to standardise the pre-test and post-test POMP scores.

**Figure 3:**
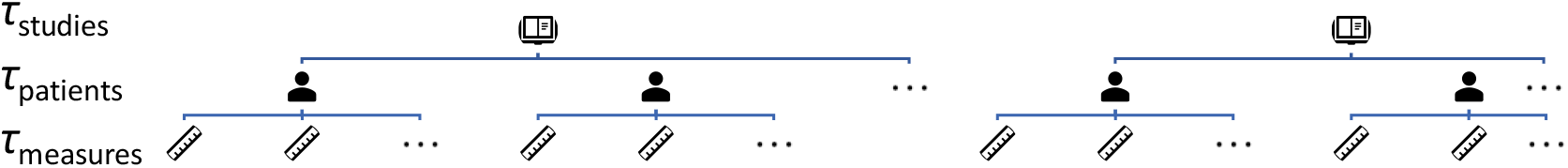
Nested multilevel model employed. Standard deviations (τ) are shown at every level: measures 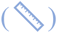, nested within patients 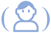, nested within studies 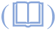.

For models specifically fitted to the RCT and the case report data respectively, see Supplementary Materials, §4.

#### 2.5.2. Moderator analyses

For the RCT meta-analyses, we fitted a meta-regression model with the moderators (1) Domain (see §2.4); (2) whether the study used validated tests for its outcome measures, or unvalidated ones (for unvalidated measures, we treated trained and untrained items as separate groups to avoid confounding measure validation and training effects); and (3) the Domain × Validated interaction. Next, to test the effect of time since stroke, we fit another model adding the additional moderators of (1) mean MPO across treatment and control groups; and (2) the difference in mean MPO between treatment and control groups.

For the case report meta-analyses, we initially fitted a similar meta-regression model with Domain, Validated, and Domain × Validated moderators. We tested additional moderators by fitting two additional models, adding one moderator at a time to this baseline model. First, we fit a model adding individual-level MPO. Second, we fit a model adding whether a study used the original MIT protocol or a modified MIT protocol.

### 2.6. Data availability

All data generated during the making of this work, including raw materials, coding sheets, analysis scripts, and supplementary materials, have been uploaded to the OSF repository https://osf.io/gcjqr/.

## 3. Results

Study-level standardised mean difference scores and meta-analytic mean differences by Domain are shown in Figure 4. Full meta-regression results tables are reported in the Supplementary Materials, eTables 12–19.

**Figure 4:**
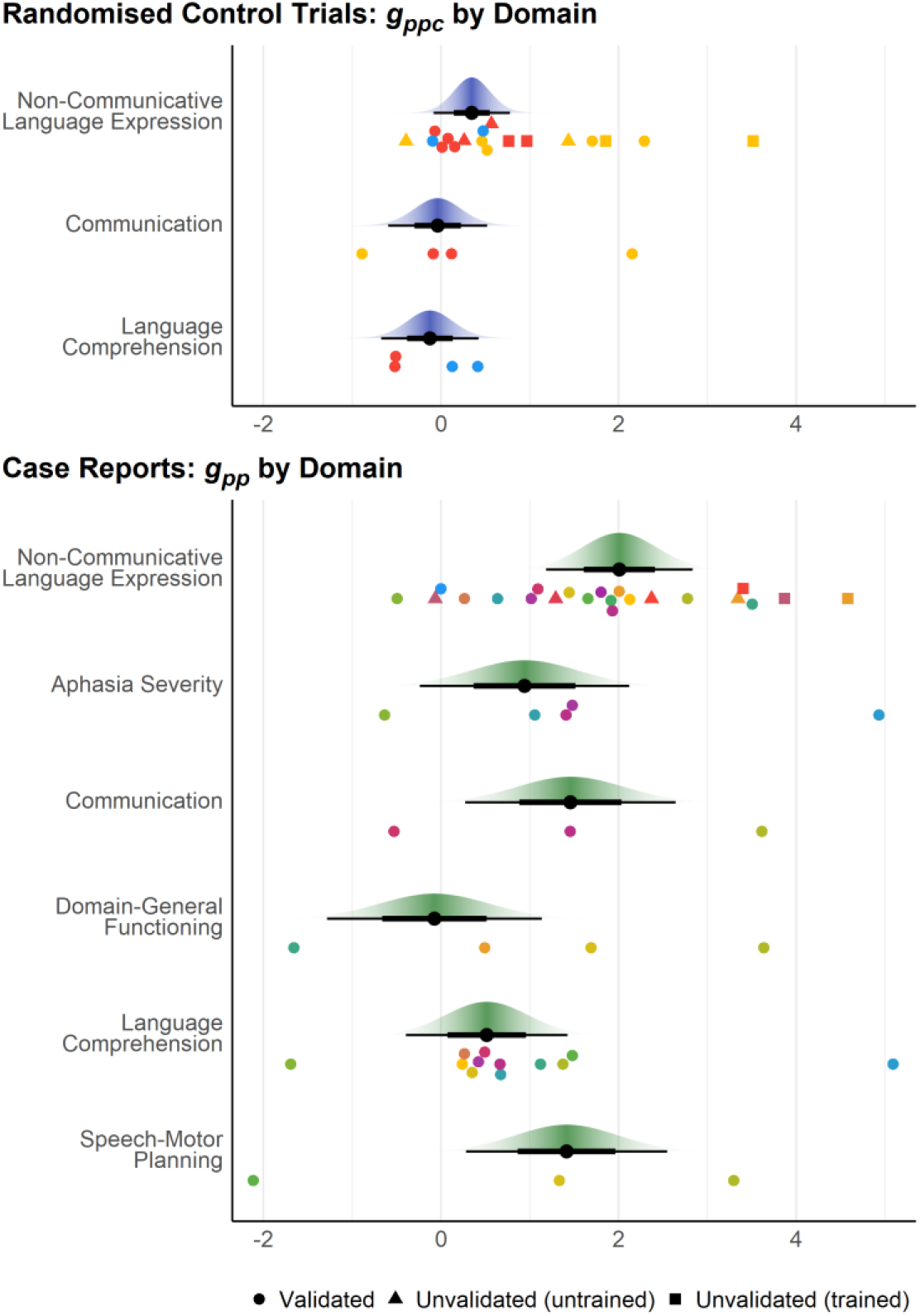
Results of meta-analyses. Data points (circles, triangles, or squares) are study-level standardised mean pretest-posttest difference scores, either adjusted for a control group (*g*_*ppc*_) or not (*g*_*pp*_). Points of different colours are drawn from different studies. Large points are mean *g*_*pp(c)*_ for validated measures with 66% (thick bar) and 95% (thin bar) confidence intervals and *t*-distribution confidence densities. For case reports, one aphasia severity study with *g*_*ppc*_ = −4.88 is not displayed.

### 3.1. RCT data

Overall, RCT data showed a small-to-moderate pretest-posttest effect of MIT on aphasia outcomes, after accounting for the control group 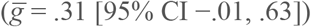. These results were primarily based on measures of Non-Communicative Language Expression (i.e., focus on verbal utterances *per se*, such as in tasks requiring repetition of words and sentences; *k* = 3, *n*_*treat*_ = 176, *n*_*control*_ = 188^b^). Other abilities were less commonly assessed, including Communication (i.e., verbal utterances used for social interaction in everyday situations; *k* = 2, *n*_*treat*_ = 39, *n*_*control*_ = 42) and Language Comprehension (i.e., understanding the meaning of verbal utterances; *k* = 2, *n*_*treat*_ = 36, *n*_*control*_ = 37). In moderator analyses, effects appeared to be much weaker for Communication and Language Comprehension tasks than for Non-Communicative Language Expression, but confidence intervals for these differences were wide (see Figure 4). Effects were estimated to be somewhat heterogeneous across studies (random effects standard deviation, *τ =* .33 [95% CI .15, 1.01]).

Two included RCTs included several unvalidated measures of Non-Communicative Language Expression. For these unvalidated measures, treatment effects for untrained items were somewhat smaller than those for trained items, though the confidence interval for this difference was fairly wide 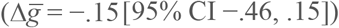. As expected, estimated treatment effects were much larger when patients were tested using trained items (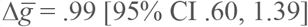]; trained vs. untrained items contrast: 1.15 [95% CI .74, 1.56]). Since measurement error tends to attenuate effect size,^27–29^ the smaller effect sizes for unvalidated measures (*k* = 2, *n*_*treat*_ = 39, *n*_*control*_ = 42) may be attributable to poorer reliability compared to validated measures (*k* = 3, *n*_*treat*_ = 173, *n*_*control*_ = 183).

When aphasia stage (MPO) was added to the RCT model (*k* = 3, *n*_*treat*_ = 251, *n*_*control*_ = 267), neither mean MPO across groups 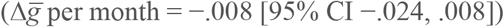 nor difference in mean MPO between MIT and control groups 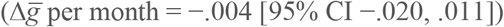 showed meaningful relationships with MIT treatment effects. Importantly, effect sizes for RCT analyses were drawn from only three studies, so these group-level MPO analyses have limited power to estimate the impact of MPO on MIT treatment effects.

### 3.2. Case report data

Compared to RCT studies, case reports with no control group estimated much larger effects of MIT 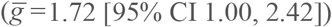. As with RCT studies, these results were primarily based on Non-Communicative Language Expression (repetition) tasks. Overall aphasia severity and language comprehension appeared to show somewhat smaller effects, but confidence intervals on these differences were very wide. Effects were estimated to be highly heterogeneous across studies (*τ* [between-studies] = 1.41 [95% CI .89, 2.05]), to the degree that MIT was even estimated to be harmful in a small proportion of settings: for instance, the 95% normal-theory prediction interval for Non-Communicative Language Expression ranged −0.88 to +4.90.^30^

Four case report studies included several unvalidated measures of Non-Communicative Language Expression (total *n* = 10). As with RCT studies, treatment effects for untrained items (on those unvalidated measures) appeared to be smaller than those for trained items, with a wide confidence interval 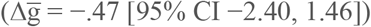. Also similar to RCTs, apparent treatment effects were much larger for trained items (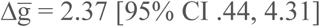; trained vs. untrained items contrast: 2.84 [95% CI 1.21, 4.48]).

When aphasia stage (MPO) was added to the case report model (*k* = 16, *n* = 246), MPO showed a moderate negative relationship with treatment effects (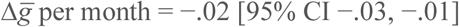; estimated effect for 12 months, −.18 [95% CI −.30, −.07]; estimated effect for 24 months, −.37 [95% CI −.61, −.14]).

Compared to studies employing the original MIT protocol (*k* = 9, *n* = 131), studies employing modified protocols (*k* = 10, *n* = 210) appeared to show somewhat larger treatment effects, though the confidence interval on this difference was very wide 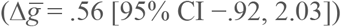.

## 4. Discussion

The present meta-analysis aimed to investigate the efficacy of MIT while accounting for crucial methodological aspects of primary studies, such as control comparisons, randomised group allocation, use of validated outcome measures, and variance in MIT protocol. It also examined the confounding effect of spontaneous recovery, and the degree to which MIT’s effect generalises to untrained items.

Overall, we found that MIT had a limited positive treatment effect in specific domains, mainly repetition tasks, in line with previous meta-analyses. However, our results reveal that poor methodology may introduce substantial bias into estimated treatment effects. Concerning RCT studies of Non-Communicative Language Expression, the use of unvalidated measures for untrained items may attenuate MIT’s effect size by about 43% when compared to validated ones (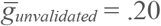 vs. 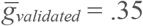). Holding language domain and outcome measure validity constant, MIT’s effect size proved to be 5.7 times larger for non-RCT data compared to RCT data (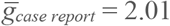 vs. 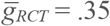 for validated Non-Communicative Language Expression measures). Implications and possible sources for each of these findings are discussed below.

### 4.1. Research implications

The current results indicate that appropriate study design can help reduce confound to obtain more realistic effect size estimates. In particular, these results re-affirm the importance of setting up and adjusting for adequate control interventions. Otherwise, most of the changes observed in case reports— evident as inflated estimates of efficacy (the 5.7 factor reported)^c^—are inseparable from phenomena of spontaneous recovery, and ultimately regression to the mean, none of which emerge from the treatment.

Figure 5 schematically illustrates the need for a control group in order to estimate the treatment effect (TE), net of influences resulting from time post-stroke only, such as impact of spontaneous recovery. Case series report T_2_-T_1_ and tend to interpret it as the TE. However, this confounds TE with the spontaneous recovery effect (SR). To isolate TE, a control group is needed: from it, we compute T_2_-C_2_ accounting for baseline differences at T_1_ to estimate the actual TE. Figure e1 in the Supplementary Discussion further illustrates this issue in the form of causal diagrams (directed acyclic graphs).

**Figure 5:**
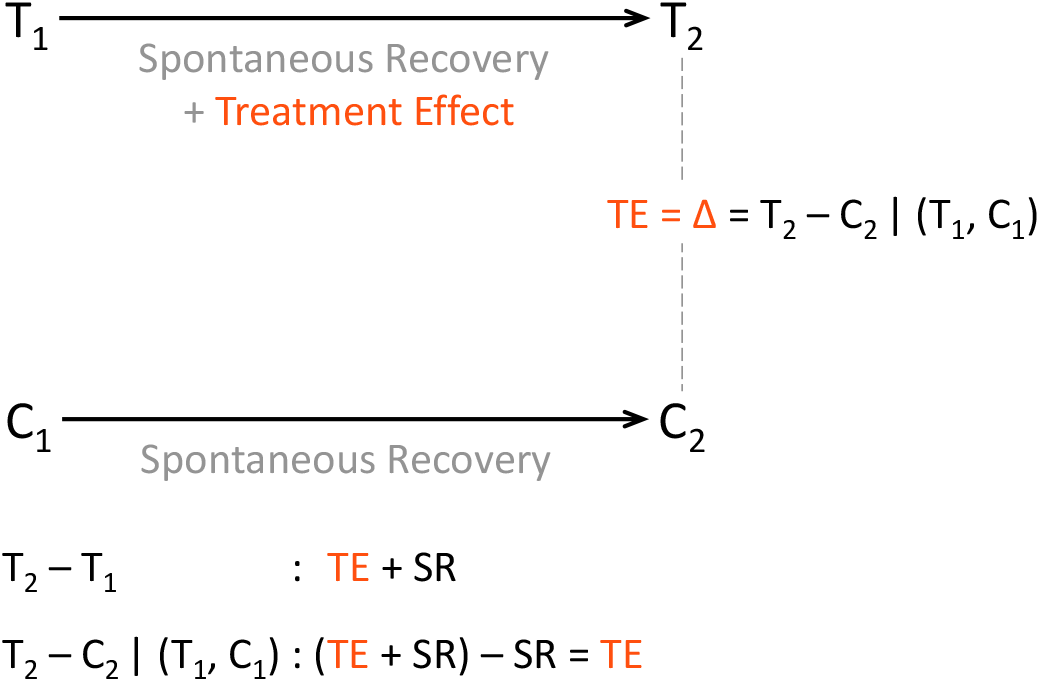
Treatment and spontaneous recovery effects in interventions. T_1_ and T_2_ represent the treatment group, at time 1 and time 2 (pre and post treatment). C_1_ and C_2_ represent the control group, at the same two time points. TE, treatment effect. The | operator denotes “accounting for”. The first equation shows how TE gets confounded with SR. The second equation shows how the confounding effect of SR can be removed. Also see Figure e1 for a causal diagram (Supplementary Materials).

Control interventions were drastically different amongst our three RCTs, namely “control therapy not aiming at language production but using linguistic tasks often trained in severe nonfluent aphasia, such as written language production, language comprehension, and nonverbal communication strategies”^9^; “no individual treatment offered, [i.e., only] social interaction as well as low intensity group therapy to support verbal and non-verbal communication”;^10^ and “none, or waiting list treatment.”^31^ Future research and its aggregation in future meta-analyses would benefit from control groups with standardised and empirically validated types of intervention, number of weekly sessions and hours of daily training.

Effect sizes were found to decrease with number of months post-stroke (MPO) for IPD studies, indicating that progress in language performance reported in the late subacute or consolidation stage of aphasia may arise from influences of spontaneous recovery^d^. Currently available data do not allow conclusions about whether MIT’s effect size increases or decreases with MPO, given the general lack of positive RCT evidence on speech-language therapy in subacute aphasia.^16^ Taken together, these results suggest that validated outcomes, randomised-controlled designs and inclusion of individuals with chronic aphasia are essential prerequisites to determine the efficacy of MIT in a reliable way.

The chronically reorganised language system post-stroke comprises undamaged perilesional tissue in the left hemisphere, as well as homotopic areas in the right hemisphere.^32^ However, the concrete distribution of activity depends on the time elapsed after stroke. In our meta-analysis, the influence of time post-onset of stroke was seen only for IPD, not for RCT studies. We submit two potential reasons for this differential finding: first, it may be a statistical artifact (fewer data points in the RCT category); second, substantial heterogeneity may exist among post-stroke recovery trajectories, which does not “subtract out” during a treatment-vs.-control comparison.^33^ The latter conjecture would require consolidation by behavioural data, possibly incorporating Saur et al.’s model of language recovery.^34^

### 4.2. Clinical implications

According to the present meta-analysis, MIT leads to gains mainly in repetition tasks that reflect the ability to reproduce prior utterances in exactly the same form. Although this ability may facilitate the acquisition of novel words, it is not entirely clear to what extent it ultimately affects verbal behaviour in everyday communicative situations.^35^ Our RCT results indicate negligible progress on validated outcomes of everyday communication ability with MIT. The number of non-repetition outcomes was comparatively small, regardless of experimental design, implying that benefits from MIT cannot be ruled out completely; nonetheless, current evidence does not support them. In contrast, large-scale RCT data demonstrate that combining selected non-MIT methods *can* lead to moderate gains on validated outcomes of communication ability.^2^ This finding suggests that individuals with aphasia should not rely exclusively on MIT if the primary goal is to improve everyday communication. Still, our meta-analysis should not be taken to downplay the importance of MIT-mediated progress on trained items. In individuals with severe forms of aphasia, a ‘palliative’ use of MIT may entail a substantial increase in quality of life.^15^ Critically, individuals with aphasia may perceive notable progress in language performance irrespective of statistically significant gains on validated outcomes. Known as ‘minimal clinically-important difference’,^36^ this diagnostic approach may be especially valuable for individuals where MIT can help establish a repertoire of trained phrases to convey basic needs in daily life.^37^ Conversely, it would be recommendable for future studies to address the impact of MIT on quality of life, emotional well-being, and severity of post-stroke depression in individuals with neurological communication disorders.^38^

### 4.3. Limitations and future directions

As with any meta-analysis, the conclusiveness of the results depends on the quantity and quality of the available sources. Our rigorous eligibility criteria left us with a relatively low number of included studies. This small sample size in turn led to large confidence intervals, which necessarily limit the strength of the conclusions. As new clinical studies using MIT become available, future meta-analyses will be able to draw conclusions and make recommendations with greater confidence, provided that subsequent studies overcome methodological issues of the sort we have pointed out.

As it stands, such issues in our included studies call for caution in interpreting the results. Our meta-analysis considered various methodological aspects largely neglected in previous work. In particular, it carefully determined the psychometric quality of each outcome measure, relative to recently defined standards in aphasia research.^39^ In addition, our evaluation accounted for quality of the research design, in terms of using control interventions and group randomisation to address unspecific influences, including bias due to placebo effects. Our results confirm the overall efficacy of MIT in repetition tasks, albeit to a smaller degree than previously reported.

Interestingly, deviation from the original MIT protocol did not systematically alter the effect size. This finding casts doubt on the notion that the original composition and hierarchical structure of MIT are indispensable for improving language performance. Although a number of studies included in our meta-analysis employed a modified MIT protocol, their individual effects are heterogeneous. Therefore, our results cannot express certainty about the impact of MIT protocol variation, and instead highlight the need for high-quality research exploring the influence of specific modifications.

Based on unvalidated outcomes, cross-sectional and longitudinal multiple-case studies have examined the role of different MIT elements: melody and rhythm,^40^ vocal expression in unison or alone,^41^ left-hand tapping,^42^ and formulaicity of verbal utterances.^43^ Possible methodological reasons for seemingly contradictory data, as well as conjectured mechanisms of MIT, have been discussed.^44^ Obviously, the present results do not offer insight into any of these mechanisms. If indeed adherence to the original MIT protocol does not manifest in significantly elevated language performance, our results encourage future research to optimise the composition and structure of the treatment to increase its efficacy in the rehabilitation of neurological communication disorders. For example, individuals with apraxia of speech may benefit from several elements of MIT, such as rhythmic pacing^45^ and language formulaicity.^46^ For now, the assumed benefit of MIT in individuals with impaired speech-motor planning remains a highly plausible hypothesis, one that our meta-analysis cannot properly address given the lack of RCT data for the mentioned patient population (*n*=8 patients across *k*=2 IPD studies). However, we believe that pursuing this particular avenue in future work may be extremely promising.

### 4.4. Conclusion

We here present the first meta-analysis on MIT that attempts to monitor the effects of various methodological caveats in interpreting the outcome of previous studies, such as lack of validated outcomes, control group or randomisation. Accounting for each of these issues in a rigorous way, the results of our meta-analyses confirm the promising role of MIT in improving language performance on trained and untrained items as well as in repetition tasks, while highlighting possible limitations in promoting everyday communication ability. We hope that the current work will help clinicians, patients and families make realistic decisions about their treatment options in the context of MIT.

## Data Availability

All data relevant for this work is present either in the main manuscript, or in the supplementary files that accompany it.

https://osf.io/gcjqr/

## 6. Acknowledgements

We gratefully acknowledge Ajay Halai and Yina Quique Buitrago for their helpful comments; Joost Hurkmans for providing unpublished work; and Sarah Wallace, Luisa Krein and Emily Braun for providing test-related information. Author TP accepts responsibility for the integrity of the data analysed.

## 7. Author contributions

CRediT author statement:

- Conceptualisation: TP, BS, WTF
- Methodology: TP, BMW
- Software: TP, BMW
- Validation: TP
- Formal analysis: TP, BMW
- Investigation: TP
- Resources: TP, BS, WTF
- Literature search and curation: TP, HH, MZ
- Data curation: TP, BMW, FH, TM
- Writing - original draft: TP, BS
- Writing - review & editing: TP, BS, BMW, FH
- Visualisation: TP, BMW
- Supervision: TP, BS
- Project administration: TP
- Funding acquisition: RB, WTF

## Footnotes

a For a small number of studies, it was not possible to determine the maximum or minimum possible scores. For these studies, we computed POMP scores using the maximum and minimum *observed* scores in the sample. Results did not change meaningfully if we excluded these studies from results.

b *k* = number of studies, *n* = number of patients. For a complete list of the number of cases (studies and patients) entering into each type of analysis, please see §5.1 of the Supplementary Materials (eTables 5 through 11).

c See also eTable 11 in the Supplementary Materials, which reports RCT meta-analyses only considering the change in control groups.

d See also eTable 12 in the Supplementary Materials, which reports IPD meta-analyses with MPO as a moderator for pretest scores only.

