## Supplementary Materials for "Melodic Intonation Therapy for aphasia: A multi-level meta-analysis of randomised controlled trials and individual participant data"

(Popescu, Stahl, et al., 2022)

*Annals of the NY Academy of Sciences*

### 1. LITERATURE SEARCH PROCEDURE

The systematic literature search in literature databases yielded 606 hits; through searching the trial registers 7 additional trials were found. These 613 items have been subjected to a duplicate check for identical publications found through the different search tools combined. They were checked with the "duplicate detection" feature of the reference management software Zotero (Corporation for Digital Scholarship; https://www.zotero.org/), with the following procedure: When all of the duplicate items had the same publication form and all of the duplicates had an abstract, we kept the first one in the list (by order of importing into Zotero); if not, we kept the first item of the duplicates having an abstract. With different publication entries from the same study, we followed the preference rule: journal article > book chapter > conference proceedings. After this process of eliminating all supplicates 143 items remained. Following this first step we read and checked all abstracts of the remaining articles against the exclusion criteria. Within this step 44 articles were excluded, leaving 99 articles that checked all eligibility criteria. As the final step the full texts of all remaining articles were reviewed. During two rounds, 78 articles were excluded, leaving 22 final articles that fit all our inclusion criteria and could therefore be included in this meta-analysis.

#### 1.1. Electronic searches

We searched the following databases: *Cochrane Library* (last searched 06.08.2021), *CINAHL EBSCOhost* (06.08.2021), *PsycINFO OVID* (1806 to August 2021), *Web of Science* (06.08.2021), *PubMed* (06.08.2021), *Scopus* (06.08.2021), *Medline OVID* (1946 to August 2021) *PSYNDEX OVID* (1977 to August 2021), *Music Periodicals Database* (06.08.2021), and *ProQuest Dissertations & Theses Global* (06.08.2021).

We also searched trial registers, including *International Clinical Trials Registry Platform* (ICTRP, https://www.isrctn.com/; 06.08.2021), *National Research Register* (UK), http://www.nihr.ac.uk/; 06.08.2021), *Clinical Trials.gov* (www.clinicaltrials.gov; 06.08.2021), *Netherlands Trials Register* www.trialregister.nl; 06.08.2021), and the *German clinical trials Register* https://www.drks.de/drks_web/; 06.08.2021)

Additionally, we performed searches in Google Scholar (06.08.2021) and in the grey literature database OpenGrey.eu (http://www.opengrey.eu/; 06.08.2021). Messages soliciting any unpublished data were additionally sent to:

- aphasia associations
  - National Aphasia Association, NAA, https://www.aphasia.org
  - Australian Aphasia Association (AAA), https://aphasia.org.au
  - Fédération Nationale des Aphasiques de France (FNAF), http://aphasie.fr/
- music therapy associations
  - American Music Therapy Association (AMTA), https://www.musictherapy.org/
  - British Association for Music Therapy (BAMT), https://www.bamt.org
- mailing lists
  - AUDITORY
  - Musicology-all
- authors of all included studies
- authors of studies for which essential data was missing; when no clarification was obtained, study was excluded (see note of eTable 2).

Finally, to ensure no studies were omitted we consulted the list of studies in published systematic reviews and meta-analyses concerning MIT [1–5]. Since no filters relating to methods used or publication type were applied to our searches, we manually separated and kept the empirical studies from the overall results. The number of records identified from each database was as follows:

- CINAHL EBSCOhost (26)
- Cochrane Library (26)
- Google Scholar (100)
- Medline OVID (48)
- Music Periodicals Database (27)
- ProQuest Dissertations & Theses Global (10)
- PsycINFO OVID (87)
- PSYNDEX OVID (6)
- PubMed (52)
- Scopus (94)
- Web of Science (130)
- Registers (n = 7)
- ClinicalTrials.gov (3)
- German Clinical Trials Register (1)
- International Clinical Trials Registry Platform (1)
- Netherlands Trials Register (2)

#### 1.2. Search terms

*CINAHL:*

((MM "Aphasia+") OR "aphasia" OR (MH "Aphasia, Broca") OR (MH "Aphasia, Transcortical Sensory") OR (MH "Aphasia, Wernicke")) AND (((MH singing or singing ) AND ("speech therapy" or (MH "Speech Therapy+")) OR "melodic intonation therapy")))

*ClinicalTrials.gov*

"melodic intonation therapy"

*Cochrane Library*

"melodic intonation therapy" AND aphasia

("speech therapy“ in Ti Abstr Key OR MeSH descriptor [speech therapy] explode all trees) AND singing* in Ti Abstr Key

*Deutsches Register klinischer Studien (DRKS)/German clinical trials register* https://www.drks.de/drks_web/

melodic intonation therapy

melodische intonationstherapie

*Google Scholar*

„melodic intonation therapy“

„melodische intonationstherapie“

*ICTRP (International Clinical Trials Registry Platform)*

"melodic intonation therapy"

*Medline OVID*

("melodic intonation therapy".ab. or "melodic intonation therapy".ti. or "melodic intonation therapy".id. or ((singing.ab. or singing.id. or singing.ti. or singing/) AND (speech therapy/ or "speech therapy".ab. or "speech therapy".id. or "speech therapy".ti.))) AND (aphasia.ab. or aphasia.id. or aphasia.ti. or exp aphasia)

*Music Periodicals Database*

"melodic intonation therapy" and aphasia

*National Research Register (UK): http://www.nihr.ac.uk/*

"melodic intonation therapy"

*Netherlands Trials Register www.trialregister.nl*

Melodic intonation

*OpenGrey.eu http://www.opengrey.eu/*

„melodic intonation therapy“

*ProQuest Dissertations & Theses Global*

„melodic intonation therapy“

*PsycINFO OVID*

("melodic intonation therapy".ab. or "melodic intonation therapy".ti. or "melodic intonation therapy".id. or ((singing.ab. or singing.id. or singing.ti. or singing/) AND (speech therapy/ or "speech therapy".ab. or "speech therapy".id. or "speech therapy".ti.))) AND (aphasia.ab. or aphasia.id. or aphasia.ti. or exp aphasia)

*PSYNDEX OVID*

("melodic intonation therapy".ab. or "melodic intonation therapy".ti. or "melodic intonation therapy".id. or ((singing.ab. or singing.id. or singing.ti. or singing/) AND (speech therapy/ or "speech therapy".ab. or "speech therapy".id. or "speech therapy".ti.))) AND (aphasia.ab. or aphasia.id. or aphasia.ti. or exp aphasia)

*PubMed*

(„melodic intonation therapy“ OR ((singing [MeSH Terms] OR singing) AND ("speech therapy"[MeSH Terms] OR "speech therapy")) AND (aphasia[MeSH Terms] OR aphasia)

*Scopus*

((( TITLE-ABS-KEY ( singing ) AND (TITLE-ABS-KEY ( "speech therapy" OR "language therapy" ))) OR ( TITLE-ABS-KEY ( "melodic intonation therapy" ))) AND ( TITLE-ABS-KEY ( aphasia))

*Web of Science*

TOPIC: (("melodic intonation therapy") OR TOPIC: ("singing" AND ("speech therapy" OR "language therapy")) AND TOPIC: aphasia

### 2. Lists of primary studies considered

#### 2.1. Included studies

eTable 1: List of studies included in the present meta-analysis.

| | Study type | First author | Year | Title | $N_{MIT}$ | $N_{ctrl}$ | IPD | MIT protocol |
|---|---|---|---|---|---|---|---|---|
| 1 | case series | Akanuma | 2016 | Singing can improve speech function in aphasics associated with intact right basal ganglia and preserve right temporal glucose metabolism: Implications for singing therapy indication | 10 | - | 1 | singing therapy |
| 2 | case series | Belin | 1996 | Recovery from nonfluent aphasia after melodic intonation therapy: A PET study | 7 | - | 1 | TMR |
| 4 | case series | Cortese | 2015 | Rehabilitation of aphasia: application of melodic-rhythmic therapy to Italian language | 6 | - | 1 | MRT |
| 5 | case series | Haro-Martínez | 2017 | Adaptation of melodic intonation therapy to Spanish: a feasibility pilot study | 4 | - | 1 | MIT |
| 9 | case series | Hurkmans | 2015 | The effectiveness of Speech–Music Therapy for Aphasia (SMTA) in five speakers with Apraxia of Speech and aphasia | 5 | - | 1 | SMTA |
| 10 | case series | Jungblut | 2014 | Paving the Way for Speech: Voice-Training-Induced Plasticity in Chronic Aphasia and Apraxia of Speech—Three Single Cases | 3 | - | 1 | SIPARI |
| 11 | case series | Naeser | 1985 | CT Scan Lesion Localization and Response to Melodic Intonation Therapy with Nonfluent Aphasia Cases | 8 | - | 1 | MIT |
| 17 | case series | van de Sandt-Koenderman | 2018 | Language lateralisation after Melodic Intonation Therapy: an fMRI study in subacute and chronic aphasia | 9 | - | 1 | MIT |
| 18 | case series | van der Meulen | 2012 | Melodic Intonation Therapy: Present Controversies and Future Opportunities | 2 | - | 1 | MIT |
| 22 | case series | Zumbansen | 2014 | The Combination of Rhythm and Pitch Can Account for the Beneficial Effect of Melodic Intonation Therapy on Connected Speech Improvements in Broca's Aphasia | 3 | - | 1 | MIT |
| 14 | controlled before&after study | Stahl | 2013 | How to engage the right brain hemisphere in aphasics without even singing: evidence for two paths of speech recovery | 5 | - | 1 | singing therapy |
| 6 | RCT | Haro-Martínez | 2019 | Melodic intonation therapy in post-stroke nonfluent aphasia: a randomized pilot trial | 20 | 20 | 0 | MIT |
| 19 | RCT | van der Meulen | 2014 | The Efficacy and Timing of Melodic Intonation Therapy in Subacute Aphasia | 23 | 25 | 0 | MIT |
| 20 | RCT | van der Meulen | 2016 | Melodic Intonation Therapy in Chronic Aphasia: Evidence from a Pilot Randomized Controlled Trial | 16 | 17 | 0 | MIT |
| 3 | single-case study | Bitan | 2018 | Changes in Resting-State Connectivity following Melody-Based Therapy in a Patient with Aphasia | 1 | - | 1 | MMIT |
| 7 | single-case study | Hatayama | 2021 | Music intonation therapy is effective for speech output in a patient with non-fluent aphasia in a chronic stage | 1 | - | 1 | MIT |
| 8 | single-case study | Homan | 2015 | A Combination of Therapeutic Techniques: Severe Broca's Aphasia | 1 | - | 1 | MMIT |
| 12 | single-case study | Primassin | 2014 | Melodische Intonationstherapie bei einer aphasischen Patientin in der (Post-) Akutphase | 1 | - | 1 | MIT |
| 13 | single-case study | Slavin | 2018 | A Case Study Using a Multimodal Approach to Melodic Intonation Therapy | 1 | - | 1 | SMTA |
| 15 | single-case study | Tabei | 2016 | Improved Neural Processing Efficiency in a Chronic Aphasia Patient Following Melodic Intonation Therapy: A Neuropsychological and Functional MRI Study | 1 | - | 1 | MIT |
| 16 | single-case study | van de Sandt-Koenderman | 2010 | A Case Study of Melodic Intonation Therapy (MIT) in the Subacute Stage of Aphasia: Early Re-re activation of Left Hemisphere Structures | 1 | - | 1 | MIT |
| 21 | single-case study | Wilson | 2006 | Preserved singing in aphasia: a case study of the efficacy of the Melodic Intonation Therapy | 1 | - | 1 | palliative MIT (pMIT) |

*Notes*. $N_{MIT}$, number of patients in MIT (treatment) group. $N_{ctrl}$, number of patients in control group. IPD, individual participant data reported.

##### 2.2. Excluded studies

eTable 2: List of initially considered, later excluded studies (also see lists of *Eligibility criteria* in main manuscript).

| Study | | | | Reason for exclusion | | | | | | |
|---|---|---|---|---|---|---|---|---|---|---|
| **Type** | **First author** | **Year** | **Title** | a | b | c | d | e | f | g |
| case series | Al-Janabi | 2014 | Augmenting melodic intonation therapy with non-invasive brain stimulation to treat impaired left-hemisphere function: two case studies | | ✓ | ✓ | ✓ | | | |
| case series | Baker | 2000 | Modifying the Melodic Intonation Therapy Program for Adults With Severe Non-fluent Aphasia | | ✓ | | | | | |
| case series | Bonakdarpour | 2003 | Melodic intonation therapy in Persian aphasic patients | | ✓ | ✓ | ✓[4] | | | |
| case series | Breier | 2010 | Changes in maps of language activity activation following melodic intonation therapy using magnetoencephalography: Two case studies | | ✓ | ✓ | | | | |
| case series | Darland | 2021 | The Effects of Varying Melodic Intervals in Melodic Intonation Therapy for Persons with Aphasia | ✓ | | | | | | |
| case series | Hurkmans | 2016 | The treatment of apraxia of speech: Speech and music therapy, an innovative joint effort | ✓[5] | | | | | | |
| case series | Kim | 2008 | Protocol Evaluation for Effective Music Therapy for Persons with Nonfluent Aphasia | | ✓ | ✓ | ✓ | | | |
| case series | Lim | 2013 | The Therapeutic Effect of Neurologic Music Therapy and Speech Language Therapy in Post-Stroke Aphasic Patients | | | | ✓ | ✓ | | |
| case series | Mauszycki | 2016 | Melodic intonation therapy applied to the production of questions in aphasia | | ✓ | ✓ | | | | |
| case series | Schlaug | 2009 | Evidence for plasticity in white-matter tracts of patients with chronic Broca’s aphasia undergoing intense intonation-based speech therapy. | | ✓ | | | | | |
| case series | Schlaug | 2008 | From Singing to Speaking: Why Singing May Lead to Recovery of Expressive Language Function in Patients with Broca's Aphasia | | ✓ | ✓ | | | | |
| case series | Sparks | 1974 | Aphasia rehabilitation resulting from melodic intonation therapy | | | | | ✓[1] | | |
| case series | Tonkovich | 1977 | The Effects of Stress and Melodic Intonation on Apraxia of Speech | | ✓ | | ✓ | | | |
| case series | Wambaugh | 2012 | Acquired Apraxia of Speech: The Effects of Repeated Practice and Rate/Rhythm Control Treatments on Sound Production Accuracy | ✓ | | | | | | |
| controlled before&after study | Osisanya | 2012 | Effectiveness of melodic intonation therapy in the management of communication difficulty of pupils with non-fluent aphasia in the classroom setting | | | | | | ✓ | |
| controlled before&after study | Wan | 2014 | Intensive therapy induces contralateral white matter changes in chronic stroke patients with Broca’s aphasia | | ✓ | | | | | |
| cross-over trial | Brendel | 2008 | Effectiveness of metrical pacing in the treatment of apraxia of speech | ✓ | | | | | | |
| cross-over trial | Krauss | 1982 | Melodic intonation therapy with language delayed apraxic children | | | | | | | ✓ |
| cross-over trial | Springer | 1993 | Training in the use of wh-questions and prepositions in dialogues: A comparison of two different approaches in aphasia therapy | | ✓[6] | | | ✓[7] | | |
| RCT | Conklyn | 2012 | The Effects of Modified Melodic Intonation Therapy on Nonfluent Aphasia: A Pilot Study | | ✓ | ✓ | | | | |

| Study | | | | Reason for exclusion | | | | | | |
|---|---|---|---|---|---|---|---|---|---|---|
| **Type** | **First author** | **Year** | **Title** | a | b | c | d | e | f | g |
| RCT | Raglio | 2016 | Improvement of spontaneous language in stroke patients with chronic aphasia treated with music therapy: a randomized controlled trial | ✓ | | | | | | |
| RCT | Vines | 2011 | Non invasive brain stimulation enhances the effects of melodic intonation therapy | ✓ | ✓ | ✓ | | | | |
| RCT | Zumbansen | 2017 | Effect of choir activity in the rehabilitation of aphasia: a blind, randomised, controlled pilot study | ✓ | | | | | | |
| single-case study | Fountura | 2014 | Efficacy of the Adapted Melodic Intonation Therapy: a case study of a Broca`s Aphasia Patient | | | | | | ✓ | |
| single-case study | Goldfarb | 1979 | Espousing melodic intonation therapy in aphasia rehabilitation: a case study. | | ✓ | ✓ | ✓[3] | | | |
| single-case study | Hough | 2010 | Melodic Intonation Therapy and aphasia: another variation on a theme | | ✓ | | | | | |
| single-case study | Jungblut | 2009 | Long-term recovery from chronic global aphasia: A case report | | | | ✓ | | | |
| single-case study | Keith | 1975 | Singing as therapy for apraxia of speech and aphasia: Report of a case | ✓ | | | | | | |
| single-case study | Lagasse | 2012 | Evaluation of Melodic Intonation Therapy for Developmental Apraxia of Speech | | | | | ✓[2] | | |
| single-case study | Marshall | 1976 | Melodic Intonation Therapy: Variations on a Theme | | | ✓ | | | | |
| single-case study | Martzoukou | 2021 | Adaptation of Melodic Intonation Therapy to Greek: A Clinical Study in Broca's Aphasia With Brain Perfusion SPECT Validation | ✓ | | | | | | |
| single-case study | Mauszycki | 2008 | The effects of rate control treatment on consonant production accuracy in mild apraxia of speech | ✓ | | | | | | |
| single-case study | Morrow-Odom | 2013 | Effectiveness of melodic intonation therapy in a case of aphasia following right hemisphere stroke | | ✓ | | | | | |
| single-case study | Wambaugh | 2000 | Effects of rate and rhythm control treatment on consonant production accuracy in apraxia of speech | ✓ | | | | | | |
| single-case study | Zipse | 2012 | When right is all that is left: plasticity of right-hemisphere tracts in a young aphasic patient | | | | | | | ✓ |

*Notes*. **a**: Substantial variation from the original MIT protocol; **b**: Non-validated tests; **c**: No contrast of trained vs. untrained items; **d**: No pre&post data; **e**: Other essential data not reported, and not retrievable even after emailing authors (post-2000 studies only); **f**: Untrustworthy source, e.g. non-peer-reviewed journal; **g**: Patient(s) 17 or younger.
[1] (i) n=2 patients were excluded who showed no improvement (publication bias); (ii) scores could not be readily converted to POMP or *z* scores. [2] SLT and MIT sessions were interleaved. [3] Unclear whether data in Fig. 3 can be taken to mean pre (Level 1) and post (Level 23). [4] Only difference data (T2-T1) reported for 'untreated variables' (all unvalidated, Table 4), which does not qualify for trained/untrained labelling. [5] SMTA (Speech–Music Therapy for Aphasia) administered in parallel with regular speech language-therapy (SLT), with no way of telling the two interventions apart. [6] AAT used for diagnosing, unclear whether the wh-questions also derived from there. [7] No between-subjects dispersion data (e.g. SEM) reported.

#### 3. TESTS AND OUTCOME MEASURES IN PRIMARY STUDIES

eTable 3: List of tests considered, across all included and excluded studies. Reference given for validation study, where identified.

| Abbreviation | Test battery full name | Validation study found |
|---|---|---|
| - | Farsi aphasia test | No |
| - | Apraxia Battery for Adults | No |
| AABT | Aachener Aphasie Bedside Test | Yes[6] |
| AAT | Aachen Aphasia Test | Yes[7,8] |
| ADP | Aphasia Diagnostic Profiles | No |
| ANELT | Amsterdam-Nijmegen Everyday Language Test | Yes[9] |
| BDAE | Boston Diagnostic Aphasia Examination | Yes[10,11] |
| BNT | Boston Naming Test | Yes[12,13] |
| CAL | Communicative Activity Log | No |
| CIU | Sabadel CIUs/min | No |
| DIAS | Diagnostic Instrument for Apraxia of Speech (AoS) | Yes[14] |
| HWL | Hierarchical Word List | Yes[15] |
| MT86 | Montréal–Toulouse Aphasia Battery | No |
| PALPA | Psycholinguistic assessments of language processing in aphasia | Yes[16] |
| PICA | Porch Index Of Communicative Ability | Yes[17,18] |
| PPTT | Pyramids and Palm Trees Test | Yes[16] |
| SLTA | Standard Language Test of Aphasia | Yes[19] |
| WAB | Western Aphasia Battery | Yes[20] |

eTable 4: Categorisation scheme showing nesting for each target syndrome: Subtests → Tests → Abilities → Domains.

| Target syndrome | Domain | Ability | Test battery | Subtest |
|---|---|---|---|---|
| Aphasia | Aphasia severity | Overall language performance | AAT | aphasia severity |
| Aphasia | Aphasia severity | Overall language performance | BDAE | aphasia severity |
| Aphasia | Aphasia severity | Overall language performance | PALPA | (no particular subtests) |
| Aphasia | Aphasia severity | Overall language performance | PPTT | (no particular subtests) |
| Aphasia | Aphasia severity | Overall language performance | WAB | aphasia quotient (AQ) |
| Aphasia | Communication | Everyday communication | ANELT | comprehensibility |
| Aphasia | Communication | Everyday communication | ANELT | intelligibility |
| Aphasia | Communication | Everyday communication | ANELT | verbal communication |
| Aphasia | Domain-general function | Cognitive-executive skills | AABT | BLIKO = Aufforderungen zu Blick- und Kopfbewegungen |
| Aphasia | Domain-general function | Cognitive-executive skills | AABT | IDENT = Identifizieren von Objekten |
| Aphasia | Domain-general function | Cognitive-executive skills | AABT | MUMO = Aufforderungen zu Mundbewegungen |
| Aphasia | Domain-general function | Cognitive-executive skills | AAT | token |
| Aphasia | Language comprehension | Auditory comprehension | AAT | auditory comprehension |
| Aphasia | Language comprehension | Auditory comprehension | BDAE | auditory commands |
| Aphasia | Language comprehension | Auditory comprehension | BDAE | auditory comprehension |
| Aphasia | Language comprehension | Auditory comprehension | BDAE | complex auditory material |
| Aphasia | Language comprehension | Auditory comprehension | WAB | auditory comprehension |
| Aphasia | Language comprehension | Written comprehension | AAT | written comprehension |
| Aphasia | Language comprehension | Written comprehension | AAT | written language |
| Aphasia | Language comprehension | Written comprehension | BDAE | reading comprehension |
| Aphasia | Language comprehension | Written comprehension | WAB | reading |
| Aphasia | Non-Comm. lang. expr. | Articulatory agility | BDAE | articulatory agility |
| Aphasia | Non-Comm. lang. expr. | Grammatical form | BDAE | grammatical form |
| Aphasia | Non-Comm. lang. expr. | Naming | AABT | BENENN = Benennen von Objekten |
| Aphasia | Non-Comm. lang. expr. | Naming | AAT | naming |
| Aphasia | Non-Comm. lang. expr. | Naming | BDAE | naming |
| Aphasia | Non-Comm. lang. expr. | Naming | BDAE | naming (confrontation ~) |
| Aphasia | Non-Comm. lang. expr. | Naming | BDAE | naming (responsive ~) |
| Aphasia | Non-Comm. lang. expr. | Naming | BNT | naming |
| Aphasia | Non-Comm. lang. expr. | Naming | SLTA | picture (manga) description test |
| Aphasia | Non-Comm. lang. expr. | Naming | WAB | naming |
| Aphasia | Non-Comm. lang. expr. | Phrase length | BDAE | phrase length |

| Target syndrome | Domain | Ability | Test battery | Subtest |
|---|---|---|---|---|
| Aphasia | Non-Comm. lang. expr. | Repetition | (trained) | repetition |
| Aphasia | Non-Comm. lang. expr. | Repetition | (untrained) | repetition |
| Aphasia | Non-Comm. lang. expr. | Repetition | AABT | SIREI = Singen, Reihen- und Floskelsprechen |
| Aphasia | Non-Comm. lang. expr. | Repetition | AAT | repetition |
| Aphasia | Non-Comm. lang. expr. | Repetition | BDAE | repetition |
| Aphasia | Non-Comm. lang. expr. | Repetition | WAB | repetition |
| Aphasia | Non-Comm. lang. expr. | Spontaneous speech | AAT | automatic language |
| Aphasia | Non-Comm. lang. expr. | Spontaneous speech | AAT | communication |
| Aphasia | Non-Comm. lang. expr. | Spontaneous speech | AAT | phonetic language |
| Aphasia | Non-Comm. lang. expr. | Spontaneous speech | AAT | prosody |
| Aphasia | Non-Comm. lang. expr. | Spontaneous speech | AAT | semantic language |
| Aphasia | Non-Comm. lang. expr. | Spontaneous speech | AAT | syntactic language |
| Aphasia | Non-Comm. lang. expr. | Spontaneous speech | WAB | spontaneous speech |
| Aphasia | Non-Comm. lang. expr. | Syllable production | (trained) | correct syllables across test-phrases |
| Aphasia | Non-Comm. lang. expr. | Syllable production | (trained) | correct syllables per test-phrase |
| Aphasia | Non-Comm. lang. expr. | Syllable production | (untrained) | correct syllables across test-phrases |
| Aphasia | Non-Comm. lang. expr. | Syllable production | (untrained) | correct syllables per test-phrase |
| Aphasia | Non-Comm. lang. expr. | Verbal expression | BDAE | verbal expression |
| Aphasia | Non-Comm. lang. expr. | Word production | (trained) | proportion of words correct |
| Aphasia | Non-Comm. lang. expr. | Word production | (untrained) | proportion of words correct |
| Aphasia | Non-Comm. lang. expr. | Writing | WAB | writing |
| Apraxia of Speech | Speech-motor planning | Speech-motor planning | DIAS | articulation of phonemes |
| Apraxia of Speech | Speech-motor planning | Speech-motor planning | DIAS | articulation of words |
| Apraxia of Speech | Speech-motor planning | Speech-motor planning | DIAS | diadochokinesis (DDK) |
| Apraxia of Speech | Speech-motor planning | Speech-motor planning | HWL | number of assessable items |
| Apraxia of Speech | Speech-motor planning | Speech-motor planning | HWL | phonemic structure |
| Apraxia of Speech | Speech-motor planning | Speech-motor planning | HWL | phonetic structure |
| Apraxia of Speech | Speech-motor planning | Speech-motor planning | HWL | speech fluency |

*Notes*. Please also see Figure 2 of the main manuscript, which depicts the same categorisation scheme in the form of a hierarchical diagram.
Non-Comm. lang. expr = Non-Communicative language expression.

### 4. SUPPLEMENTARY META-ANALYSIS METHODS

#### 4.1. RCT data

All RCTs were reported at the group-level. We computed effect sizes as the pretest-posttest-control group Hedges' $g$:[21]

$$g_{ppc} = \left(z_{treat_{post}} - z_{treat_{pre}}\right) - \left(z_{contr_{post}} - z_{contr_{pre}}\right).$$

We computed the variance for each $g$ using the method of Morris (2008). We estimated multi-level mixed effects meta-regression models to account for effect size dependency, with random intercepts for each study. We first fit an overall meta-analysis combining all effect sizes. Second, we fit additional meta-regression models including potential moderator variables. For these meta-regression models, we included random slopes for the *Domain* moderator, nested within studies.[22] We used a homoscedastic compound symmetric structure for the random effects, estimating a single random effects variance and correlation for all abilities.[a] We estimated the amount of heterogeneity (i.e., $\tau^{\tau}$ ) using the restricted maximum-likelihood estimator.[23] We computed confidence intervals for meta-regression coefficients and mean treatment effects using the Knapp and Hartung *t*-distribution method,[24] and for the random effects components using profile likelihood. We estimated models using *R* version 4.1.0[25] and the *metafor* package version 3.-01[b].[26]

#### 4.2. Case report data

All case reports reported results as individual-level data, so we analysed these studies using IPD meta-analysis. We computed individual-level scores as the difference between pre-test and post-test *z*-scores (the mean difference in these scores is the pretest-posttest Hedges' $g$, $g_{pp}g_{pp}$). We then pooled data across studies using a three-level random-effects IPD meta-analysis, with individual scores again (see Figure 2 in main article) nested within patients nested within studies.[27] Similar to the group-level RCT meta-analyses, we first fit an overall model including all data points with no moderators, then fit additional models including potential moderator variables as predictors. For these models, we included random intercepts for patients and studies.[c] We estimated random effects components using REML and computed confidence intervals using profile likelihood. We estimated models using *R*[25] and the *lme4* package version 1.1-27.[28]

### 5. COMPLETE RESULTS TABLES

#### 5.1. Number of cases entering into the analyses

The tables below report the total number of cases (studies and patients) entering the various analyses, broken down by various factors.

eTable 5: Number of cases for IPD studies, grouped by Domain. *k* = number of studies, *n* = number of patients.

| | *k* | *n* |
|---|---|---|
| Aphasia Severity | 6 | 21 |
| Communication | 3 | 21 |
| Domain-General Functioning | 4 | 17 |
| Language Comprehension | 12 | 62 |
| Non-Communicative Language Expression | 19 | 192 |
| Speech-Motor Planning | 3 | 28 |

[a] For comparison, we also estimated models with unequal random effects variances across dependent variables. This did not improve model fit based on AICc comparison or likelihood ratio tests.
[b] As only three RCT studies were identified, it was not possible to apply methods to detect publication-bias or other small-sample effects (e.g., tests of funnel plot asymmetry).
[c] Models with random slopes for the *Domain* variable did not converge, likely due to the limited co-occurrence of specific pairs of those within any one study.

eTable 6: Number of cases for IPD studies, grouped by outcome measure (validated/unvalidated) and test items (trained/untrained). *k* = number of studies, *n* = number of patients.

| | k | n |
|---|---|---|
| Unvalidated measures \| trained items | 4 | 10 |
| Unvalidated measures \| untrained items | 4 | 10 |
| Validated measures | 16 | 321 |

eTable 7: Number of cases for IPD studies, grouped by MIT protocol (original/modified). *k* = number of studies, *n* = number of patients.

| | *k* | *n* |
|---|---|---|
| Modified MIT | 10 | 210 |
| Original MIT | 9 | 131 |

eTable 8: Number of cases for IPD studies for which MPO data was available (patient level). *k* = number of studies, *n* = number of patients.

| | *k* | *n* |
|---|---|---|
| MPO data available | 16 | 246 |

eTable 9: Number of cases for RCT studies, grouped by Domain.

| | $k_{studies}$ | $k_{es}$ | $n_{treat}$ | $n_{control}$ |
|---|---|---|---|---|
| Communication | 2 | 4 | 39 | 42 |
| Language Comprehension | 2 | 4 | 36 | 37 |
| Non-Communicative Language Expression | 3 | 18 | 176 | 188 |

*Notes*. $k_{studies}$ = number of studies; $k_{es}$ = number of effect sizes reported across studies; $n_{treat}$ = number of patients in "treatment" groups; $n_{control}$ = number of patients in "control" groups.

eTable 10: Number of cases for RCT studies, grouped by outcome measure (validated/unvalidated) and test items (trained/untrained).

| | $k_{studies}$ | $k_{es}$ | $n_{treat}$ | $n_{control}$ |
|---|---|---|---|---|
| Unvalidated measures \| trained items | 2 | 4 | 39 | 42 |
| Unvalidated measures \| untrained items | 2 | 4 | 39 | 42 |
| Validated measures | 3 | 18 | 173 | 183 |

*Notes*. $k_{studies}$ = number of studies; $k_{es}$ = number of effect sizes reported across studies; $n_{treat}$ = number of patients in "treatment" groups; $n_{control}$ = number of patients in "control" groups.

eTable 11: Number of cases for RCT studies for which MPO data was available (at group level).

| | $k_{studies}$ | $k_{es}$ | $n_{treat}$ | $n_{control}$ |
|---|---|---|---|---|
| MPO data available | 3 | 26 | 251 | 267 |

*Notes*. $k_{studies}$ = number of studies; $k_{es}$ = number of effect sizes reported across studies; $n_{treat}$ = number of patients in "treatment" groups; $n_{control}$ = number of patients in "control" groups.

#### 5.2. RCT data

##### 5.2.1 Overall RCT meta-analyses

eTable 12: Overall RCT meta-analyses.

| Term | Estimate | *SE* | Statistic | *df* | *p* | 95% conf. int. |
|---|---|---|---|---|---|---|
| $\bar{g}$ | 0.31 | 0.16 | 2.00 | 25 | 0.057 | [−0.01, 0.63] |
| τ | 0.25 | | 212.08 | 25 | < 0.001 | [ 0.10, 1.11] |

*Notes.* $\bar{g}$ = mean pretest-posttest difference ($g_{ppc}$; accounting for control group), where a value of $\bar{g}$=1 can be back-transformed to approx. 10 points on a combined ANELT scale, based on means and SDs computed for the total ANELT norm sample;[9,29,30] τ = estimated random effects standard deviation across studies; Statistic = *t* value for $\bar{g}$ and $Q_E$ value for τ; confidence intervals computed using *t* distributions for $\bar{g}$ and profile likelihood for τ and ρ.

##### 5.2.2 RCT meta-analyses of domain categories

eTable 13: RCT meta-analyses of domain categories.

| Term | Estimate | *SE* | Statistic | *df* | *p* | 95% conf. int. |
|---|---|---|---|---|---|---|
| $\bar{g}$ (Non-Communicative Language Expression) | 0.35 | 0.21 | 1.68 | 21 | 0.108 | [-0.08, 0.78] |
| $\bar{g}$ (Communication) | -0.04 | 0.27 | -0.14 | 21 | 0.893 | [-0.59, 0.52] |
| $\bar{g}$ (Language Comprehension) | -0.12 | 0.26 | -0.47 | 21 | 0.643 | [-0.67, 0.42] |
| $\Delta\bar{g}$ (unvalidated measure with untrained items) | -0.15 | 0.15 | -1.06 | 21 | 0.300 | [-0.46, 0.15] |
| $\Delta\bar{g}$ (unvalidated measure with trained items) | 0.99 | 0.19 | 5.24 | 21 | < .001 | [ 0.60, 1.39] |
| τ | 0.33 | | 158.71 | 21 | < .001 | [ 0.15, 1.01] |
| ρ | -0.05 | | | | | [-0.52, 0.93] |

*Notes.* $\bar{g}$ = mean pretest-posttest difference ($g_{ppc}$; accounting for control group), where a value of $\bar{g}$=1 can be back-transformed to approx. 10 points on a combined ANELT scale, based on means and SDs computed for the total ANELT norm sample;[9,29,30] τ = estimated random effects standard deviation across studies; ρ = estimated correlation among *g* treatment effects between measures of different ability domains across studies; Statistic = *t* value for $\bar{g}$ and $Q_E$ value for τ; confidence intervals computed using *t* distributions for $\bar{g}$ and profile likelihood for τ.

##### 5.2.3 RCT meta-analyses with only the change in control groups

eTable 14: Meta-analyses with only the change in control groups (taken from the RCTs).

| Term | Estimate | *SE* | Statistic | *df* | *p* | 95% conf. int. |
|---|---|---|---|---|---|---|
| $\bar{g}$ (Non-Communicative Language Expression) | 0.35 | 0.23 | 1.53 | NA | 0.14 | [-0.12, 0.81] |
| $\bar{g}$ (Communication) | 0.33 | 0.24 | 1.37 | NA | 0.19 | [-0.17, 0.82] |
| $\bar{g}$ (Language Comprehension) | 0.38 | 0.23 | 1.61 | NA | 0.12 | [-0.11, 0.86] |
| $\Delta\bar{g}$ (unvalidated measure with untrained items) | 0.03 | 0.11 | 0.31 | NA | 0.76 | [-0.20, 0.26] |
| $\Delta\bar{g}$ (unvalidated measure with trained items) | -0.55 | 0.10 | -5.59 | NA | < .001 | [-0.75, -0.34] |
| τ | 0.38 | | 113.00 | 21.00 | < .001 | [ 0.17, 1.61] |
| ρ | 1.00 | | | | | [ 0.14, 1.00] |

*Notes*. Table shows that the estimated change for control groups is about .35 across categories. This accounts for some but not all of the difference in results between the $\bar{g}$ values from the RCT meta-analysis of pretest-posttest-control group, and those of the case series.

#### 5.3. Case report data (IPD)

##### 5.3.1 Overall IPD meta-analyses

eTable 15: Overall IPD meta-analyses.

| Term | Estimate | *SE* | *t* | 95% conf. int. |
|---|---|---|---|---|
| $\bar{g}$ | 1.72 | 0.35 | 4.91 | [1.00, 2.42] |
| τ | 1.25 | | | [0.75, 1.90] |
| σ (person) | 0.75 | | | [0.35, 1.16] |
| σ (measure) | 2.02 | | | [1.87, 2.20] |

*Notes.* $\bar{g}$ = mean pretest-posttest difference ($g_{pc}$; not accounting for any control group), where a value of $\bar{g}$=1 can be back-transformed to approx. 10 points on a combined ANELT scale, based on means and SDs computed for the total ANELT norm sample;[9,29,30] τ = estimated random effects standard deviation across studies; σ (person) = estimated random effects standard deviation across persons (within study); σ (measure) = estimated random effects standard deviation across measures (within person); confidence intervals computed using profile likelihood; *p* values omitted as the appropriate denominator degrees of freedom for linear mixed effects models is ill-defined;[31,32] inference should be based on the profile likelihood confidence intervals.

##### 5.3.2 IPD meta-analyses of domain categories

eTable 16: IPD meta-analyses of domain categories.

| Term | Estimate | *SE* | *t* | 95% conf. int. |
|---|---|---|---|---|
| $\bar{g}$ (Non-Communicative Language Expression) | 2.01 | 0.42 | 4.79 | [ 1.17, 2.82] |
| $\bar{g}$ (Aphasia Severity) | 0.94 | 0.60 | 1.57 | [-0.23, 2.11] |
| $\bar{g}$ (Communication) | 1.46 | 0.60 | 2.42 | [ 0.29, 2.63] |
| $\bar{g}$ (Domain-General Function) | -0.07 | 0.61 | -0.12 | [-1.27, 1.12] |
| $\bar{g}$ (Language Comprehension) | 0.52 | 0.46 | 1.11 | [-0.40, 1.41] |
| $\bar{g}$ (Speech-Motor Planning) | 1.42 | 0.58 | 2.46 | [ 0.29, 2.53] |
| $\Delta\bar{g}$ (unvalidated measure with untrained items) | -0.47 | 0.99 | -0.48 | [-2.40, 1.46] |
| $\Delta\bar{g}$ (unvalidated measure with trained items) | 2.37 | 0.99 | 2.38 | [ 0.44, 4.31] |
| τ | 1.41 | | | [ 0.89, 2.05] |
| σ (person) | 0.82 | | | [ 0.49, 1.20] |
| σ (measure) | 1.86 | | | [ 1.70, 2.01] |

*Notes.* $\bar{g}$ = mean pretest-posttest difference ($g_{pc}$; not accounting for any control group), where a value of $\bar{g}$=1 can be back-transformed to approx. 10 points on a combined ANELT scale, based on means and SDs computed for the total ANELT norm sample;[9,29,30] $\Delta\bar{g}$ = estimated difference in $\bar{g}$ between validated and unvalidated measures; note that only Non-Communicative Language Expression included unvalidated measures; τ = estimated random effects standard deviation across studies; σ (person) = estimated random effects standard deviation across persons (within study); σ (measure) = estimated random effects standard deviation across measures (within person); confidence intervals computed using profile likelihood; *p* values omitted as the appropriate denominator degrees of freedom for linear mixed effects models is ill-defined;[31,32] inference should be based on the profile likelihood confidence intervals.

##### 5.3.3 IPD meta-analyses with aphasia stage (MPO) as a moderator

eTable 17: IPD meta-analyses with aphasia stage (months post-onset, MPO) as a moderator.

| Term | Estimate | *SE* | *t* | 95% conf. int. |
|---|---|---|---|---|
| $\overline{g}$ (Non-Communicative Language Expression) | 1.97 | 0.29 | 6.74 | [ 1.39, 2.54] |
| $\overline{g}$ (Aphasia Severity) | 1.08 | 0.51 | 2.13 | [ 0.14, 2.07] |
| $\overline{g}$ (Communication) | 2.10 | 0.45 | 4.62 | [ 1.21, 3.05] |
| $\overline{g}$ (Domain-General Function) | 2.00 | 0.56 | 3.58 | [ 0.96, 3.15] |
| $\overline{g}$ (Language Comprehension) | 0.74 | 0.35 | 2.10 | [ 0.05, 1.43] |
| $\overline{g}$ (Speech-Motor Planning) | 1.96 | 0.41 | 4.74 | [ 1.14, 2.83] |
| $\Delta\overline{g}$ (unvalidated measure with untrained items) | -0.14 | 0.65 | -0.22 | [-1.41, 1.07] |
| $\Delta\overline{g}$ (unvalidated measure with trained items) | 2.70 | 0.65 | 4.15 | [ 1.43, 3.91] |
| $\Delta\overline{g}$ (per month post-onset) | -0.02 | 5.00e-03 | -3.07 | [-0.03, -0.01] |
| τ | 0.32 | | | [ 0.00, 0.82] |
| σ (person) | 1.01 | | | [ 0.69, 1.38] |
| σ (measure) | 1.57 | | | [ 1.40, 1.71] |

*Notes*. $\overline{g}$ = mean pretest-posttest difference ($g_{pc}$; not accounting for any control group), where a value of $\overline{g}$=1 can be back-transformed to approx. 10 points on a combined ANELT scale, based on means and SDs computed for the total ANELT norm sample;[9,29,30] $\Delta\overline{g}$ = estimated difference in $\overline{g}$; note that only Non-Communicative Language Expression included unvalidated measures; τ = estimated random effects standard deviation across studies; σ (person) = estimated random effects standard deviation across persons (within study); σ (measure) = estimated random effects standard deviation across measures (within person); confidence intervals computed using profile likelihood; *p* values omitted as the appropriate denominator degrees of freedom for linear mixed effects models is ill-defined;[31,32] inference should be based on the profile likelihood confidence intervals.

##### 5.3.4 IPD meta-analyses with MIT protocol as a moderator

eTable 18: IPD meta-analyses with MIT protocol as a moderator.

| Term | Estimate | *SE* | *t* | 95% conf. int. |
|---|---|---|---|---|
| $\overline{g}$ (Non-Communicative Language Expression) | 1.71 | 0.58 | 2.93 | [ 0.59, 2.83] |
| $\overline{g}$ (Aphasia Severity) | 0.64 | 0.73 | 0.88 | [-0.75, 2.03] |
| $\overline{g}$ (Communication) | 1.16 | 0.73 | 1.60 | [-0.22, 2.55] |
| $\overline{g}$ (Domain-General Function) | -0.37 | 0.74 | -0.50 | [-1.78, 1.04] |
| $\overline{g}$ (Language Comprehension) | 0.22 | 0.62 | 0.35 | [-0.97, 1.40] |
| $\overline{g}$ (Speech-Motor Planning) | 1.11 | 0.71 | 1.55 | [-0.25, 2.47] |
| $\Delta\overline{g}$ (unvalidated measure with untrained items) | -0.50 | 1.00 | -0.50 | [-2.42, 1.40] |
| $\Delta\overline{g}$ (unvalidated measure with trained items) | 2.35 | 1.00 | 2.35 | [ 0.42, 4.25] |
| $\Delta\overline{g}$ (modified MIT protocol) | 0.56 | 0.77 | 0.73 | [-0.92, 2.03] |
| τ | 1.42 | | | [ 0.84, 2.00] |
| σ (person) | 0.82 | | | [ 0.49, 1.21] |
| σ (measure) | 1.86 | | | [ 1.70, 2.01] |

*Notes*. $\overline{g}$ = mean pretest-posttest difference ($g_{pc}$; not accounting for any control group), where a value of $\overline{g}$=1 can be back-transformed to approx. 10 points on a combined ANELT scale, based on means and SDs computed for the total ANELT norm sample;[9,29,30] $\Delta\overline{g}$ = estimated difference in $\overline{g}$; note that only Non-Communicative Language Expression included unvalidated measures; τ = estimated random effects standard deviation across studies; σ (person) = estimated random effects standard deviation across persons (within study); σ (measure) = estimated random effects standard deviation across measures (within person); confidence intervals computed using profile likelihood; *p* values omitted as the appropriate denominator degrees of freedom for linear mixed effects models is ill-defined;[31,32] inference should be based on the profile likelihood confidence intervals.

##### 5.3.5 IPD meta-analyses with aphasia stage (months post-onset, MPO) as a moderator for pretest scores only

eTable 19: IPD meta-analyses with aphasia stage (months post-onset, MPO) as a moderator for pretest scores only.

| Term | Estimate | *SE* | *t* | 95% conf. int. |
|---|---|---|---|---|
| $\bar{g}$ (Non-Communicative Language Expression) | -0.19 | 0.35 | -0.54 | [-0.86, 0.48] |
| $\bar{g}$ (Aphasia Severity) | -0.66 | 0.66 | -0.99 | [-1.90, 0.63] |
| $\bar{g}$ (Communication) | 0.55 | 0.58 | 0.94 | [-0.57, 1.67] |
| $\bar{g}$ (Domain-General Function) | -0.92 | 0.74 | -1.24 | [-2.34, 0.51] |
| $\bar{g}$ (Language Comprehension) | 2.34 | 0.50 | 4.68 | [ 1.38, 3.30] |
| $\bar{g}$ (Speech-Motor Planning) | 1.55 | 0.53 | 2.95 | [ 0.54, 2.56] |
| $\Delta\bar{g}$ (unvalidated measure with untrained items) | -0.53 | 0.84 | -0.62 | [-2.15, 1.10] |
| $\Delta\bar{g}$ (unvalidated measure with trained items) | 0.55 | 0.84 | 0.65 | [-1.08, 2.18] |
| $\Delta\bar{g}$ (per month post-onset) | 0.01 | 0.01 | 0.97 | [-0.01, 0.03] |
| $\tau$ | 1.18 | | | [ 0.75, 1.61] |
| $\sigma$ (person) | 0.00 | | | [ 0.00, 0.90] |
| $\sigma$ (measure) | 2.18 | | | [ 1.94, 2.37] |

*Notes*. Table shows that pretest scores appear to increase by about .01 SDs per month post onset.

#### 6. SUPPLEMENTARY DISCUSSION

Similar to Figure 4 in the main manuscript, Figure e1 schematically demonstrates the need for a control group in order to estimate a treatment effect (TE) that is net of any effects due merely to the passage of time, such as (in disease) spontaneous recovery. It does so by depicting the "difference among differences" in an alternative form, namely as a causal diagram, a directed acyclic graph; see e.g. Ref. 33. The figure illustrates that merely comparing scores before & after having *Received MIT*, as in case series, necessarily means that TE is confounded with *Time*. *Receiving MIT* and *Time* are however perfectly correlated (coterminous), and as such cannot be isolated. By adding a control group (as in RCTs), we can estimate the effect of *Time*, in the absence of having *Received MIT*. Namely, we subtract the Before/After difference of the Control group from the Before/After difference of the Treated group, to isolate the TE.

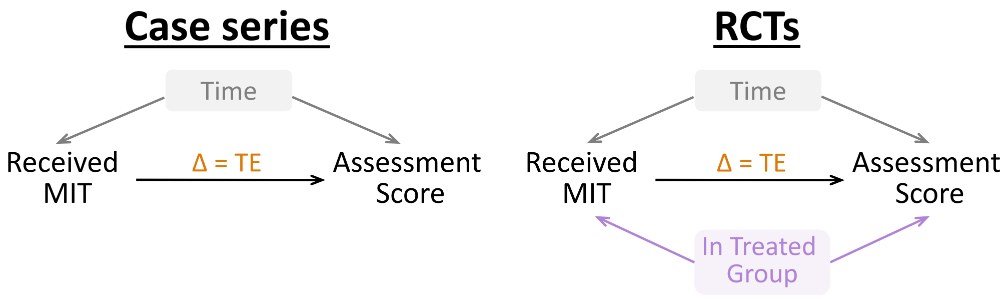

**Figure e1: Treatment and spontaneous recovery effects in interventions**, illustrated with causal diagrams (directed acyclic graphs). In case series (left-hand side), pretest-posttest differences confound treatment effects (TE) and secular time-related trends (here: spontaneous recovery). No statistical adjustment is possible to remove this confounding. In RCTs (right-hand side), the presence of a control group allows TE and spontaneous recovery to be disentangled, by comparing the condition groups (treated vs. control).
